# Gaussian graphical modeling of the serum exposome and metabolome reveals interactions between environmental chemicals and endogenous metabolites involved in critical metabolic processes

**DOI:** 10.1101/2020.09.15.20190413

**Authors:** Vincent Bessonneau, Roy R. Gerona, Jessica Trowbridge, Rachel Grashow, Thomas Lin, Heather Buren, Rachel Morello-Frosch, Ruthann A. Rudel

## Abstract

Given the complex exposures from both exogenous and endogenous sources that an individual experiences during life, exposome-wide association studies that interrogate levels of small molecules in biospecimens have been proposed for discovering causes of chronic diseases. We conducted a study to explore associations between environmental chemicals and endogenous molecules using Gaussian graphical models (GGMs) of non-targeted metabolomics data measured in a cohort of California women firefighters and office workers. GGMs revealed many exposure-metabolite associations, including that exposures to mono-hydroxyisononyl phthalate, ethyl paraben and 4-ethylbenzoic acid were associated with metabolites involved in steroid hormone biosynthesis, and perfluoroalkyl substances (PFAS) were linked to bile acids – hormones that regulate cholesterol and glucose metabolism – and inflammatory signaling molecules. Some hypotheses generated from these findings were confirmed by analysis of data from the National Health and Nutrition Examination Survey (NHANES). Taken together, our findings demonstrate a novel approach to discovering associations between chemical exposures and biological processes of potential relevance for disease causation.

---

Increasing evidence suggests that environmental, rather than genetic factors are the major causes of most chronic diseases. A recent study from Western European of monozygotic twins estimated that disease risk attributable to genetic plus shared environmental exposures ranged from 3.4% for leukemia to 48.6% for asthma with a median value of 18.5%^1^. Therefore, exploring associations between myriad exposures – originating from diet, lifestyle factors, consumer products and other sources of chemicals – received during the life course [i.e. the “exposome”^2^] and chronic diseases has the potential to elucidate new disease risk factors. Metabolomics is recognized as a powerful approach for characterizing the exposome since it can measure thousands of small molecules in biospecimens^3-5^. These small molecules can be either substrates or end products of cellular metabolism and can originate from exogenous sources via inhalation, ingestion and dermal absorption, or from endogenous processes including human and microbial metabolism. This approach has been used to discover the joint microbial/human metabolism of the nutrient choline as a potential major cause of coronary heart disease^6-8^ as well as novel risk factors for type 2 diabetes^9^, hypertension^10^, and all-cause mortality^11^ in the U.S. population.

We recently demonstrated that the application of non-targeted metabolomics using high-resolution mass spectrometry can detect more than 600 environmental chemicals in serum samples^12^. This approach can also detect both exogenous and endogenous molecules in biospecimens, making metabolomics data ideal for exploring associations between environmental exposures and metabolic changes that then generate hypotheses for targeted follow-up studies. One way to achieve this is to evaluate correlation networks between exogenous and endogenous small molecules. However, a major challenge of this method is to identify direct associations, because Pearson correlations are generally high in omics data, and so it can generate indirect associations that are not biologically relevant. One approach to circumvent the selection of indirect associations is to use Gaussian graphical models (GGMs). A GGM is an undirected probabilistic graphical model based on partial correlation coefficients between two variables (i.e. pairwise Pearson coefficients conditioned against all remaining variables)^13^. Therefore, GGMs provide an estimate of the conditional dependencies between variables (i.e. metabolites). Previous studies using GGMs have demonstrated that high partial correlations between small molecules correspond to known metabolic reactions^13-15^.

In this study, we employed GGMs to identify direct associations between environmental chemicals and endogenous metabolites measured in serum samples from 69 California women firefighters (FF) and 74 office workers (OW) enrolled in the Women Firefighters Biomonitoring Collaborative (WFBC) study^12,16^ using non-targeted liquid chromatography coupled to high-resolution mass spectrometry (LC-HRMS). Then, we used National Health and Nutrition Examination Survey (NHANES) data to test several hypotheses generated from exposure-metabolite associations and evaluate whether these exposures to environmental chemicals are associated with phenotypic responses in adult women from the general U.S. population.

## RESULTS

Non-targeted metabolomics analysis via LC-HRMS in negative ionization mode tentatively identified 145 small molecules in serum samples of women FF and OW. Most of these molecules were annotated as level 2 or 3 according to the metabolomics standard initiative^17^.

### GGM networks identify direct associations between small molecules

For this analysis, GGMs were built by combining the serum exposome data and metabolome data. The LC-HRMS data matrix contained 69 samples x 145 molecules (55 environmental chemicals and 90 endogenous metabolites), 74 samples x 139 molecules (49 environmental chemicals and 90 endogenous metabolites), and 143 samples x 142 molecules (52 environmental chemicals and 90 endogenous metabolites) for women FF, OW and the whole cohort, respectively. After applying GGM of the serum exposome and metabolome for the whole cohort, we found 74 significant partial correlations [partial correlation coefficients (PCC) ranged from −0.21 to 0.49, *p-values (p)* ranged from 2.2×10^-16^ to 1.9×10^-4^] at a False Discovery Rate (FDR) threshold of 0.1 (Fig. 1C). When stratifying by occupational group, we observed 85 (PCC ranged from −0.18 to 0.29, *p* ranged from 2.2×10^-16^ to 3.3×10^-4^) and 54 (PCC ranged from −0.17 to 0.30*, p* ranged from 1.1×10^-15^ to 1.4×10^-4^) significant partial correlations for women FF (Fig. 1A) and OW (Fig. 1B), respectively (Supplementary Table 1).

**Figure 1.**
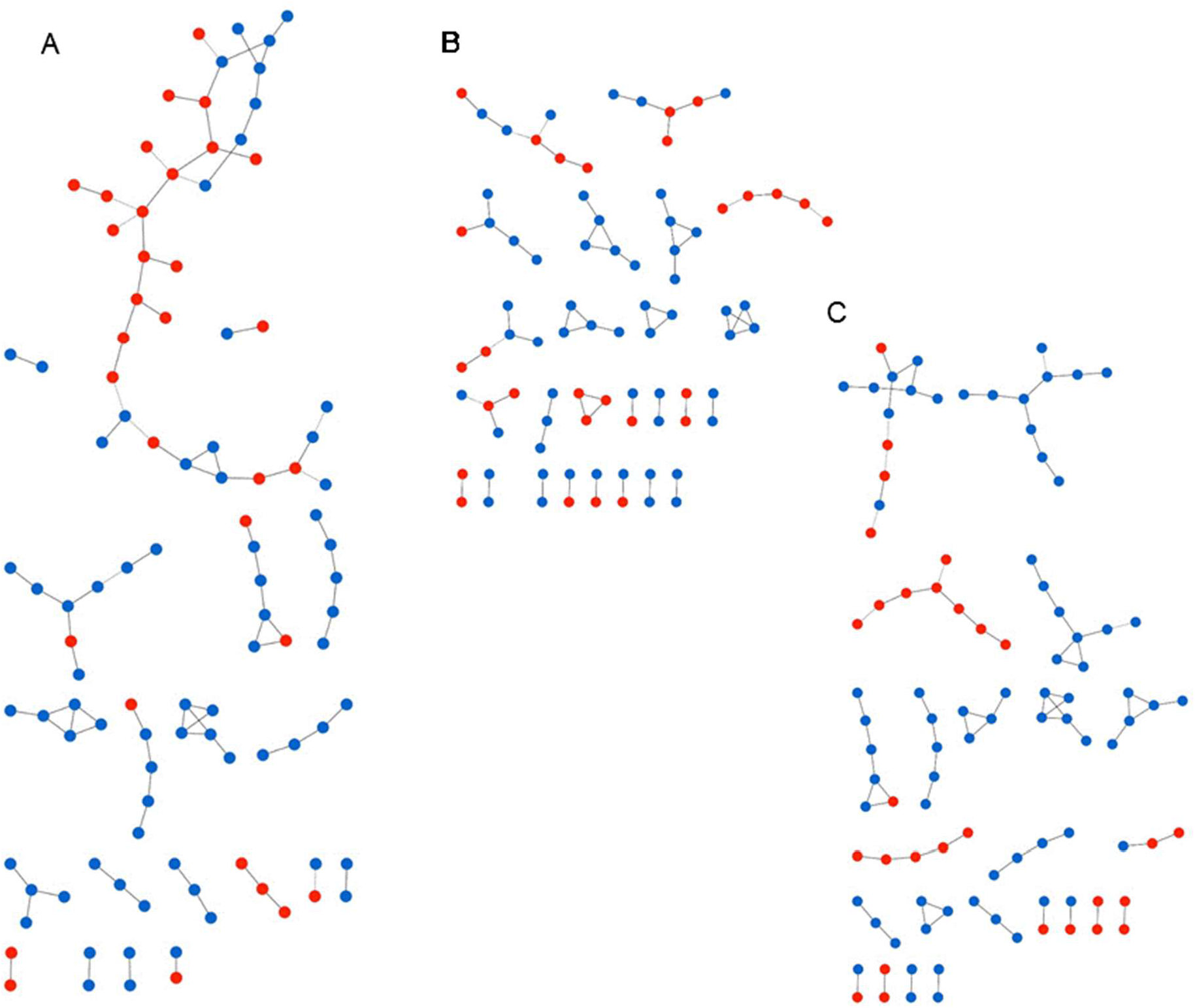
Network representation of Gaussian graphical models (GGM) of the serum exposome and metabolome measured in women firefighters (A), in women office workers (B), and in the whole cohort (C). Blue and red nodes represent endogenous metabolites and environmental chemicals, respectively. Edges connecting nodes represent significant partial correlations at FDR <10%.

### GGM reconstructs biochemical reactions

GGMs displayed numerous significant partial correlations between endogenous metabolites which were organized into sub-networks. For example, we observed strong partial correlations between several fatty acids, including linoleic acid, stearidonic acid, eicosapentaenoic acid, docosapentaenoic acid, and docosahexaenoic acid (Fig. 2A) as well as between unconjugated and conjugated bile acids (Fig. 2C) in all three GGMs (i.e. FF, OW and the whole cohort) (Supplementary Information). Using metabolic pathway databases (i.e. KEGG PATHWAY and SMPDB), we found that these molecules are in close proximity in the metabolic network related to their biosynthesis and degradation. As shown in Fig. 2B, the significant association between alpha-linoleic acid and stearidonic acid (PCC=0.20, 2×10^-6^) can be attributed to a desaturation step regulated by the FADS2 gene, while eicosapentaenoic acid is synthesized from stearidonic acid (PCC=0.19, 6×10^-6^) via ELOVL5-dependent elongation and FADS1-dependent desaturation. Similarly, we found that the significant associations between bile acids represented known biochemical reactions involved in the biosynthesis of primary and secondary bile acids via cytochrome P450-mediated oxidation of cholesterol in multi-step process^18^ (Fig. 2D). Taurodeoxycholic acid and deoxycholic acid glycine are secondary bile acids resulting from bacterial dehydroxylation in the colon of taurocholic acid (PCC=0.27, 5×10^-11^) and cholic acid glycine (PCC=0.18, 9×10^-6^), respectively^18^.

**Figure 2.**
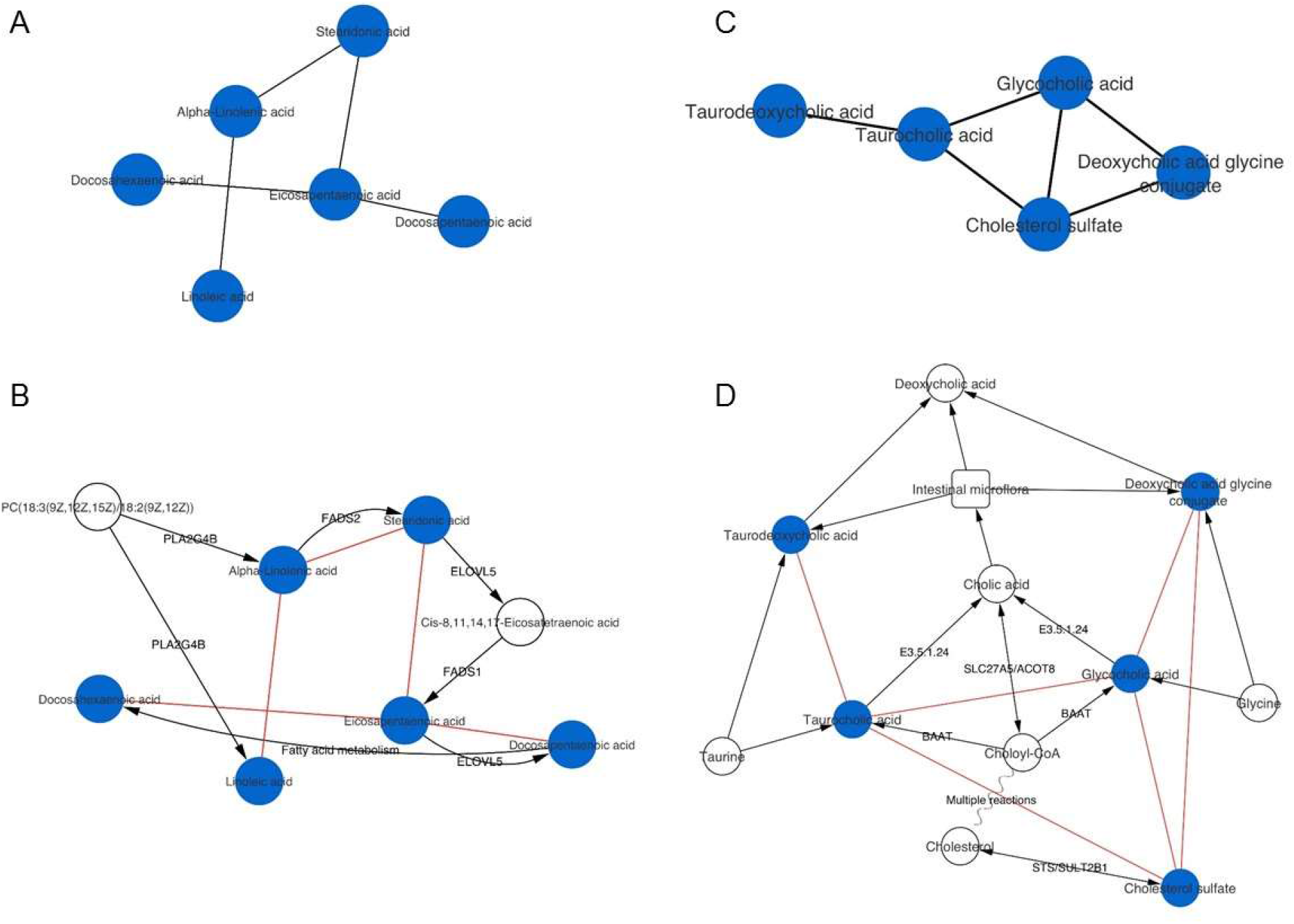
Linoleic acid (A-B) and bile acid metabolism (C-D) models inferred from GGMs of women FF and OW. A and C represent the sub-networks obtained from GGMs. B and D represent overlapping sub-networks of known biochemical reactions (black edges; labels correspond to regulating genes) obtained from KEGG pathway and from GGMs (red edges).

### Identification of exposure-metabolite associations and hypothesis generation

Among the significant partial correlations observed, we discovered many exposure-metabolite associations (Table 1). In women FF, we found that exposures to mono-hydroxyisononyl phthalate, ethyl paraben and 4-ethylbenzoic acid were associated with metabolites involved in steroid hormone biosynthesis (11β-hydroxyprogesterone and 11β-hydroxyandrosterone-3-glucuronide) (Fig. 3A). We also observed that 4-hydroxyacetophenone was negatively associated with chenodeoxycholic acid (PCC = −0.13, *p* = 1.0×10^-4^) and positively associated with lithocholic acid glycine (PCC = 0.15, *p* = 1.1×10^-5^) – bile acids involved in cholesterol homeostasis and energy balance (Fig. 3B). Exposures to perfluorohexanesulfonic acid (PFHxS) was also positively associated with one microbial-derived secondary bile acid (sulfolithocholylglycine, PCC = 0.12, *p* = 2.6×10^-5^), while perfluorooctane sulfonic acid (PFOS) was correlated with one inflammatory signaling molecule (15d PGD2, PCC = −0.13, *p* = 2.4×10^-4^) and calcitriol, the biologically active form of vitamin D (PCC = 0.14, *p* = 4.4×10^-5^) in women FF (Fig. 3B). In women OW, we observed a lower number of exposure-metabolite interactions (Table 1). We found that several phenols, including 4-hexyloxyphenol, butyl paraben, octylphenol diethoxylate and pentachlorophenol were associated with molecules involved in inflammation (12-HETE, eicosapentaenoic acid, alpha-linolenic acid and 12-HTT). We also observed that mono-isobutyl phthalate was negatively associated with two metabolites of linoleic acid metabolism and lipid peroxidation (DiHODE and 9-HODE). When combining FF and OW exposome and metabolome data, we discovered only nine associations between chemical exposures and metabolites (Table 1).

**Table 1.**
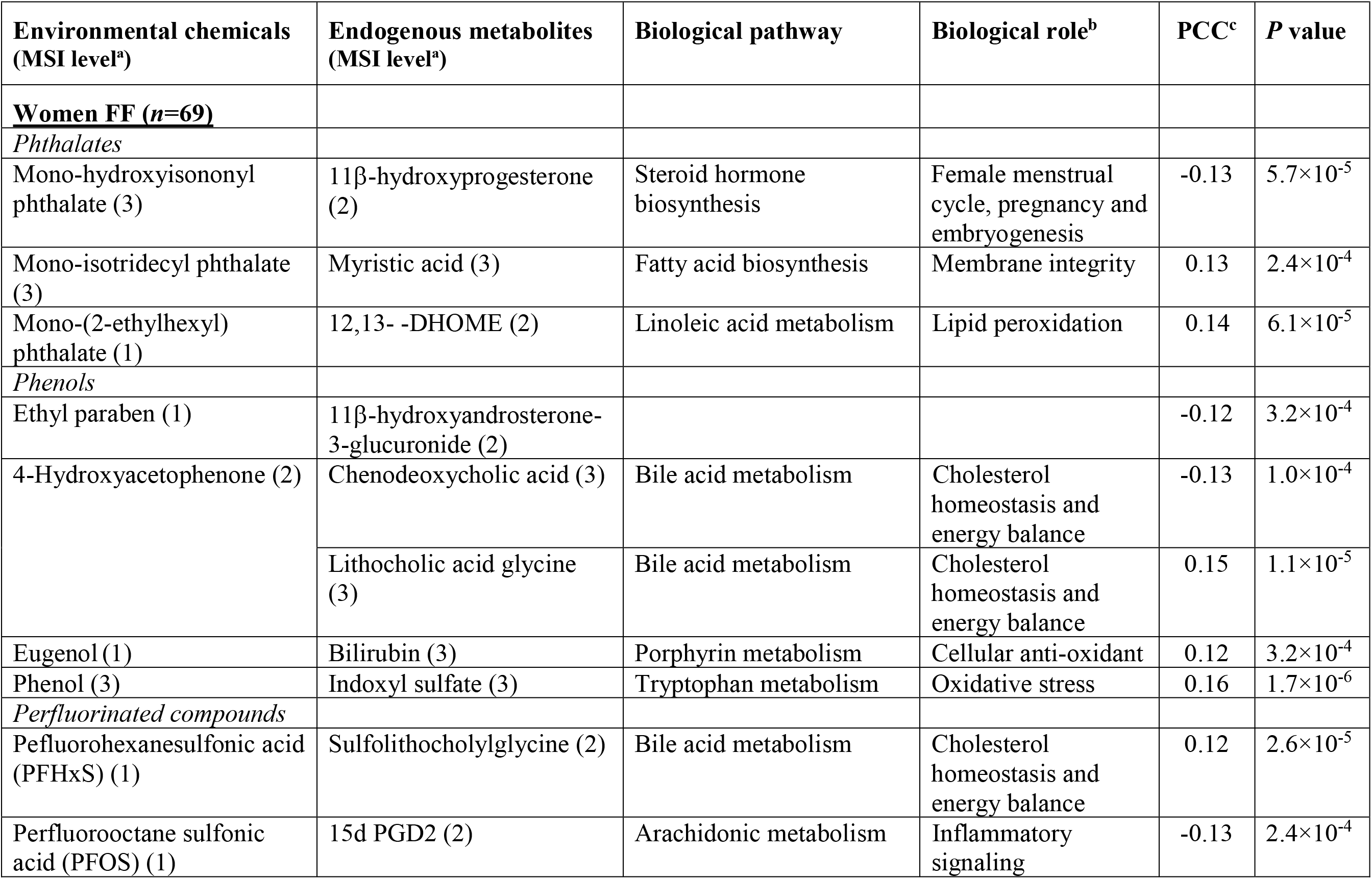
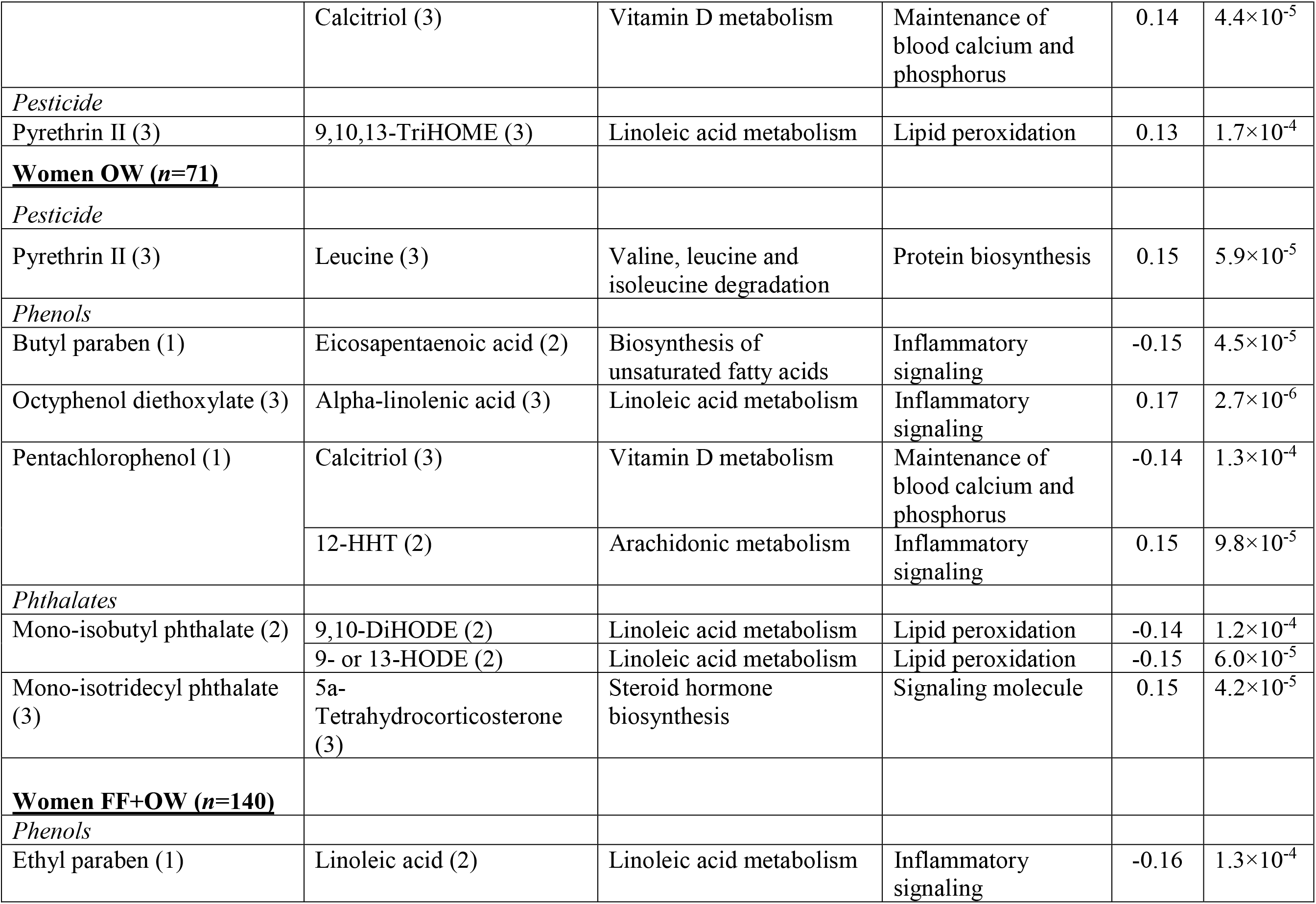
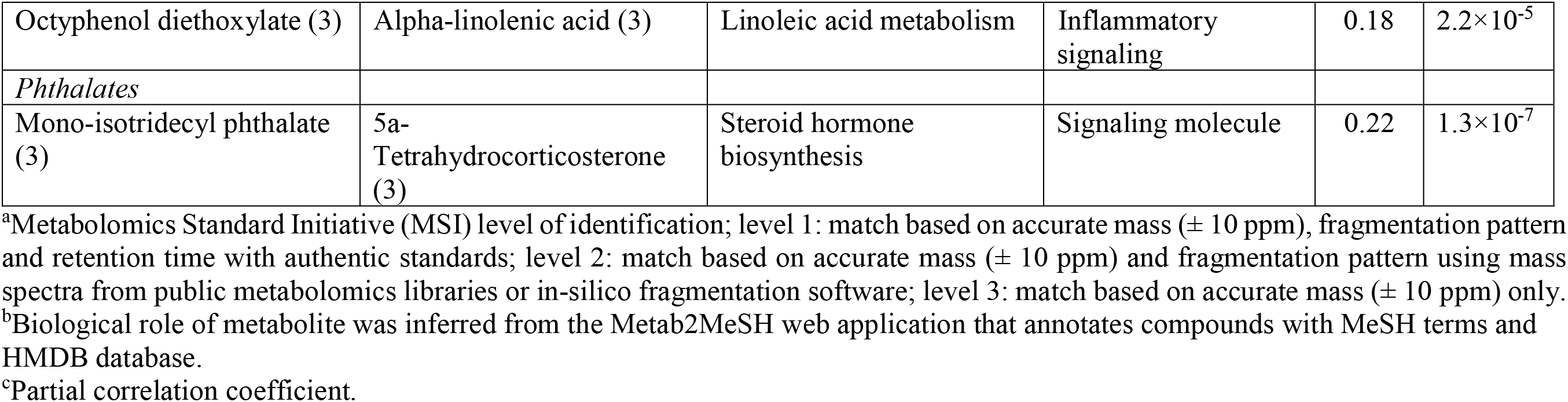
Summary of chemical-metabolite associations, biological changes and biological role of metabolites.

**Figure 3.**
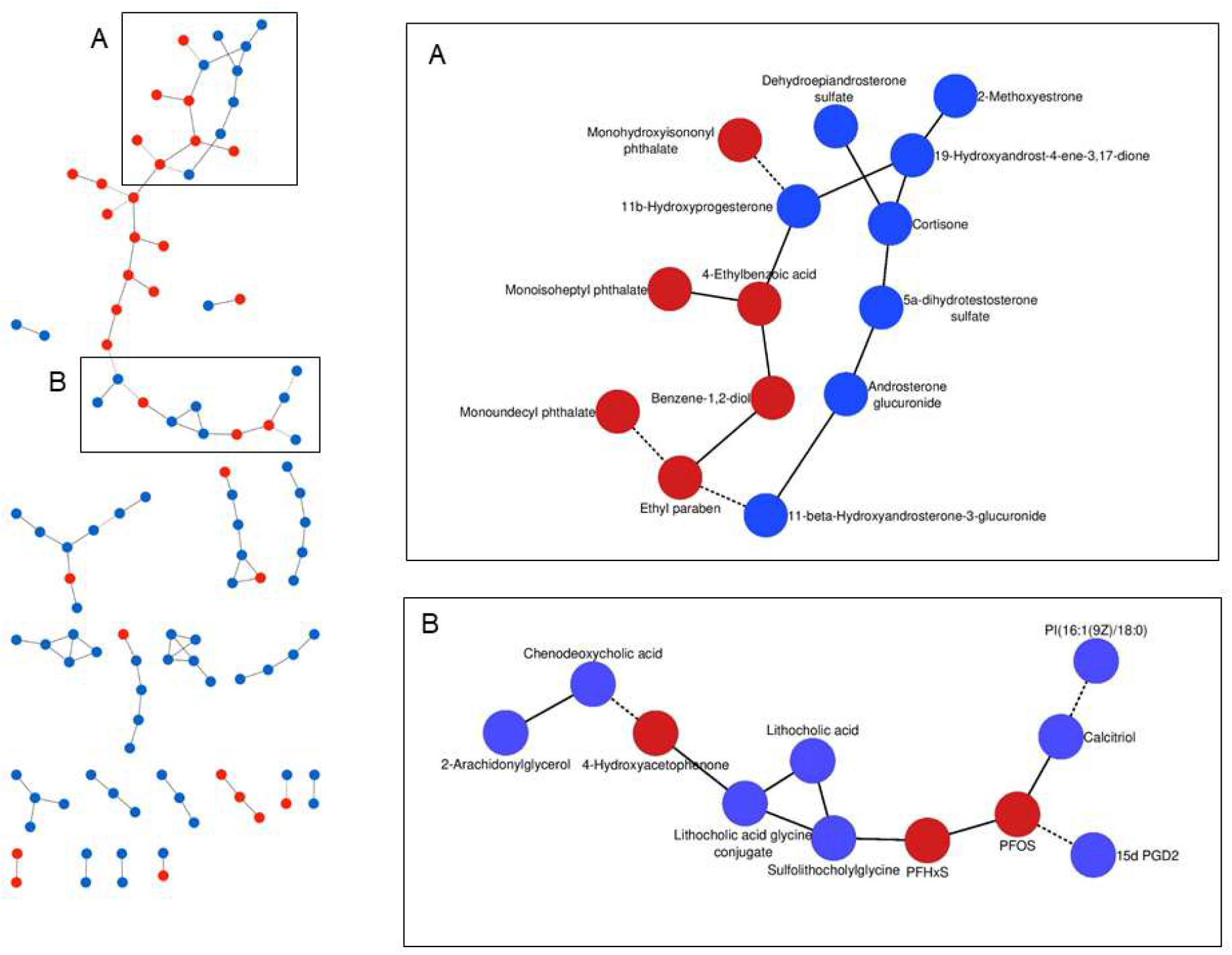
Exposure-metabolites subnetworks identified by the GGMs inferred from the women FF metabolomics dataset. The subnetwork A represents associations of phenols and phthalates with metabolites involved in steroid hormone biosynthesis. The subnetwork B shows associations between 4-hydroxyacetophenone, PFOS and PFHxS and metabolites involved bile acids, arachidonic and vitamin D metabolism. Blue and red nodes represent endogenous metabolites and environmental chemicals, respectively. Plain and dashed edges connecting nodes represent positive partial correlations and negative partial correlations, respectively.

Next, we sought to confirm exposure-metabolite associations observed from the GGM networks and evaluate whether exposure-induced metabolic perturbations are associated with phenotypic responses in adult women from the general U.S. population using NHANES. Among the significant exposure-metabolite associations, twelve were not tested because those chemicals were not measured in any NHANES cycles.

### Exposures to PFHxS and metabolic syndrome (MetS)

We observed that exposures to PFHxS were associated with changes in bile acid metabolism in women FF, and so we assessed whether exposure to this environmental chemical was linked to changes in clinical measures related to cholesterol homeostasis and energy balance in NHANES. We tested for associations between serum PFHxS concentrations and MetS and individual components of MetS (Fig. 4) in women 20-79 years of age enrolled in NHANES 2003-2014.

**Figure 4.**
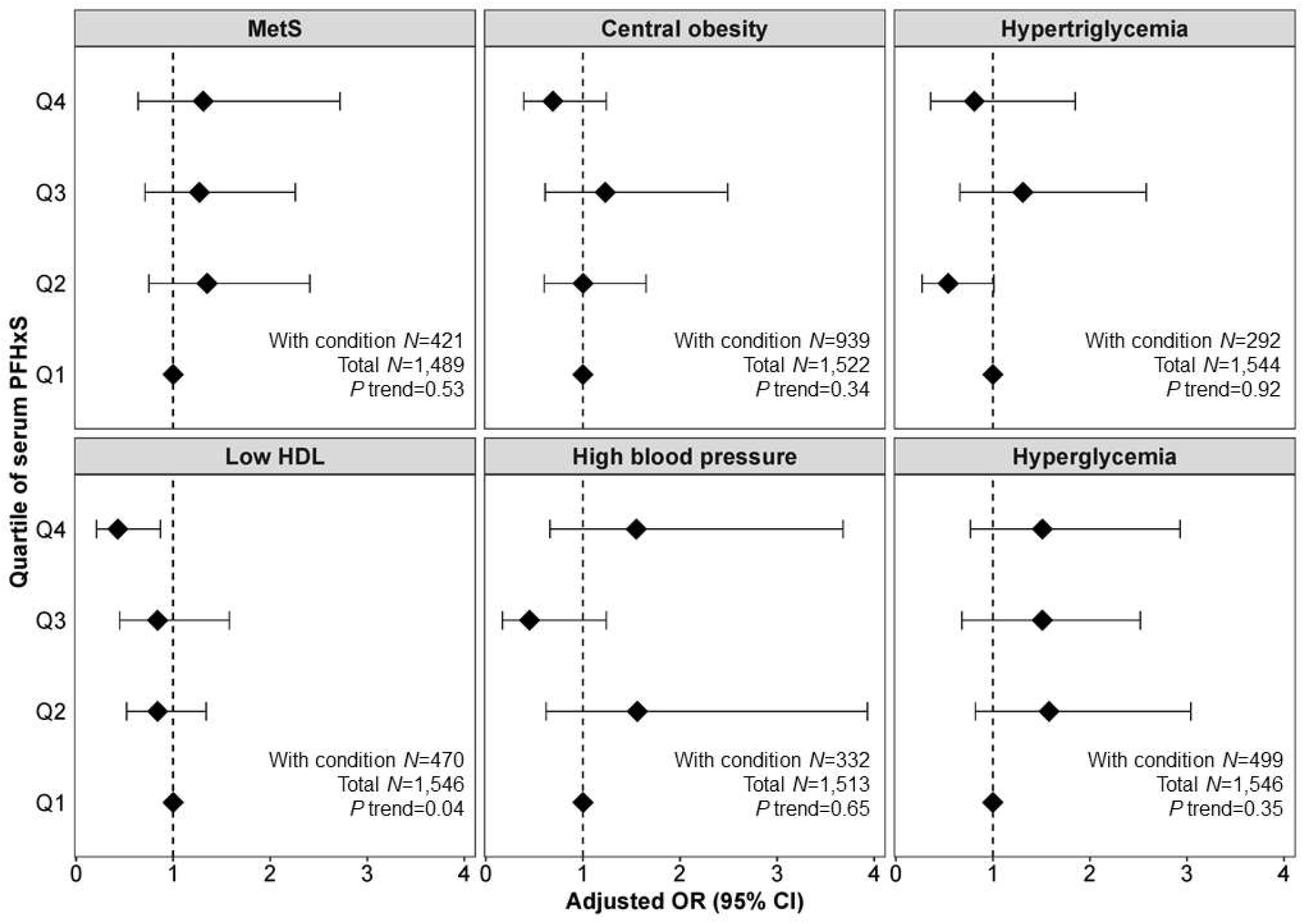
Odds ratio (95% CI) for metabolic syndrome (MetS) and individual components of MetS in women 20-79 years of age enrolled in NHANES 2003-2014 by quartile of serum PFHxS concentration. Models were adjusted for age, race/ethnicity, poverty, total caloric intake, physical activity, and smoking status.

Of the adult women in the NHANES sample, 28.3 *%* with measured PFHxS met NCEP/ATPIII criteria for MetS. No clear associations were observed for PFHxS and MetS. Since we found that exposures to this chemical were associated with a microbial-derived bile acid in our GGMs, we also tested whether the differences in gut microbiota can confound associations with MetS. However, further adjusting for recent use of antibiotics as a proxy of microbial composition had no effect on the odds ratios (ORs) for MetS (data not shown).

Weak associations were found for PFHxS and individual components of MetS, with only the association with low HDL cholesterol reaching statistical significance (adj. OR for Q4 versus Q1: 0.43; 95 *%* CI: 0.21, 0.88). Further adjusting for recent use of antibiotics had no effect on the odds ratios (ORs) for MetS components and did not change the effect estimates (data not shown).

### Exposures to PFAS and parabens and inflammatory responses

Because we observed significant partial correlations between PFOS, PFHxS, ethyl paraben and butyl paraben and inflammatory signaling molecules in women FF and OW, we tested whether these exposures are associated with clinical markers of inflammatory responses in adult women in NHANES by exploring associations between serum or urinary concentration of these chemicals and clinical markers of inflammation and immune system function (Table 2).

**Table 2.** Associations between markers of inflammation and immune system and each 100% increase in serum PFAS (NHANES 2003-2010) and urinary parabens (NHANES 2005-2010) concentrations in women 20-79 years of age enrolled in NHANES.

| | C-reactive protein<br>(mg/dL) | | White blood cells<br>(x10<sup>3</sup> cells/μL) | | Lymphocytes<br>(x10<sup>3</sup> cells/μL) | |
| --- | --- | --- | --- | --- | --- | --- |
|  | *n* | % change<sup>a</sup><br>(95% CI) | *n* | % change<sup>a</sup><br>(95% CI) | *n* | % change<sup>a</sup><br>(95% CI) |
| **Serum PFOS<sup>b</sup>** | 1,693 | **13 (4, 23)** | 2,042 | 1.2 (-0.9, 1.9) | 2,039 | **4.5 (2.1, 6.9)** |
| **Serum PFHxS** |  |  |  |  |  |  |
| Model 1<sup>b</sup> | 1,002 | -6.9 (-16, 1.8) | 1,012 | -1.6 (-3.5, 0.4) | 1,012 | 1.5 (-0.9, 3.8) |
| Model 2<sup>f</sup> | 1,002 | -6.9 (-16, 1.8) | 1,012 | -1.6 (-3.5, 0.4) | 1,012 | 1.5 (-0.8, 3.9) |
| **Urinary EPB** |  |  |  |  |  |  |
| Model 1<sup>c</sup> | 1,529 | **-7.5 (-11.7, -3.2)** | 1,556 | -0.9 (-1.9, -0.01) | 1,552 | -1.2 (-2.3, -0.02) |
| Model 2<sup>d</sup> | 1,529 | -1.7 (-5.4, 2.0) | 1,556 | -0.4 (-1.4, 0.6) | 1,552 | -0.8 (-1.9, 0.3) |
| **Urinary BPB** |  |  |  |  |  |  |
| Model 1<sup>c</sup> | 1,322 | **-6.7 (-11.3, -2.2)** | 1,472 | **-1.1 (-1.8, -0.3)** | 1,468 | -1.1 (-2.1, 0.04) |
| Model 2<sup>d</sup> | 1,322 | -1.1 (-5.4, 3.1) | 1,472 | -0.3 (-1.2, 0.4) | 1,468 | -0.6 (-1.7, 0.5) |
| | Monocytes (x10<sup>3</sup> cells/μL) | | Neutrophils (x10<sup>3</sup> cells/μL) | | Platelets (x10<sup>3</sup> cells/μL) | |
|  | *n* | % change<sup>a</sup><br>(95% CI) | *n* | % change<sup>a</sup><br>(95% CI) | *n* | % change<sup>a</sup><br>(95% CI) |
| **Serum PFOS<sup>b</sup>** | 2,039 | 1.4 (-1.6, 4.5) | 2,039 | -0.5 (-3.1, 2.1) | 2,042 | -0.5 (-1.8, 0.7) |
| **Serum PFHxS** |  |  |  |  |  |  |
| Model 1<sup>b</sup> | 1,012 | -0.5 (-3.5, 2.6) | 1,012 | **-3.5 (-5.9, -0.9)** | 1,012 | -1.9 (-4.0, 0.17) |
| Model 2<sup>f</sup> | 1,012 | -0.6 (-3.6, 2.5) | 1,012 | **-3.4 (-6.0, -0.9)** | 1,012 | -1.9 (-4.1, 0.14) |
| **Urinary EPB** |  |  |  |  |  |  |
| Model 1<sup>c</sup> | 1,552 | -0.5 (-1.3, 0.3) | 1,552 | -0.9 (-2.1, 0.4) | 1,552 | -0.1 (-1.3, 1.0) |
| Model 2<sup>d</sup> | 1,552 | -0.3 (-1.2, 0.5) | 1,552 | -0.2 (-1.5, 1.1) | 1,552 | -0.5 (-1.7, 0.7) |
| **Urinary BPB** |  |  |  |  |  |  |
| Model 1<sup>c</sup> | 1,468 | -0.2 (-1.1, 0.7) | 1,468 | -0.9 (-1.9, 0.04) | 1,472 | -0.01 (-0.8, 0.8) |
| Model 2<sup>d</sup> | 1,468 | 0.001 (-0.9, 0.9) | 1,468 | -0.1 (-1.2, 1.0) | 1,472 | 0.3 (-0.5, 1.1) |

|  | Eosinophils (x10<sup>3</sup> cells/μL) |  | Basophils (x10<sup>3</sup> cells/μL) |  |
| --- | --- | --- | --- | --- |
|  | *n* | % change<sup>a</sup><br>(95% CI) | *n* | % change<sup>a</sup><br>(95% CI) |
| **Serum PFOS<sup>b</sup>** | 2,039 | -0.2 (-1.0, 0.6) | 2,039 | 0.3 (-0.15, 0.79) |
| **Serum PFHxS** |  |  |  |  |
| Model 1<sup>b</sup> | 1,012 | -0.4 (-1.1, 0.3) | 1,012 | 0.1 (-0.3, 0.6) |
| Model 2<sup>f</sup> | 1,012 | -0.4 (-1.1, 0.3) | 1,012 | 0.1 (-0.3, 0.6) |
| **Urinary EPB** |  |  |  |  |
| Model 1<sup>c</sup> | 1,556 | -0.03 (-0.3, 0.3) | 1,556 | -0.2 (-0.4, 0.03) |
| Model 2<sup>d</sup> | 1,556 | 0.05 (-0.3, 0.4) | 1,556 | -0.1 (-0.4, 0.06) |
| **Urinary BPB** |  |  |  |  |
| Model 1<sup>c</sup> | 1,468 | -0.02 (-0.3, 0.3) | 1,468 | -0.01 (-0.3, 0.1) |
| Model 2<sup>d</sup> | 1,468 | 0.08 (-0.2, 0.4) | 1,468 | -0.01 (-0.2, 0.1) |

Association estimates were obtained with general linear models using sampling weights and accounting for the complex survey design.
<sup>a</sup>Adjusted for age, race/ethnicity, poverty, and log-transformed serum cotinine
<sup>b</sup>Adjusted for the same variables in a plus BMI
<sup>c</sup>Adjusted for the same variables in a plus log-transformed urinary creatinine
<sup>d</sup>Adjusted for the same variables in c plus BMI
<sup>e</sup>Adjusted for the same variables in c plus recent use of antibiotics
<sup>f</sup>Adjusted for the same variables in b plus recent use of antibiotics

For PFAS, we found that each 100 *%* increase in serum PFOS concentration was associated with 13 % increase in CRP (95 % CI: 4, 23) and 4.5 % increase in absolute lymphocyte count (95 % CI: 2.1, 6.9), whereas we observed a negative association between each 100 % increase in serum PFHxS concentration and absolute neutrophil count (3.5 % decrease; 95 % CI: −5.9, −0.9) after adjusting for age, race/ethnicity, poverty, BMI and serum cotinine. For parabens, we found negative associations between CRP and each 100 % increase in urinary ethyl paraben (7.5 % decrease; 95 % CI: −11.7, −3.2) and butyl paraben (6.7 % decrease; 95 % CI: −11.3, −2.2) concentrations, after adjusting for age, race/ethnicity, poverty, urinary creatinine and serum cotinine. Urinary butyl paraben was also associated with 1% decrease in white blood cell count (95 *%* CI: −1.8, −0.3). The associations between urinary parabens and CRP disappeared after further adjusting for BMI. For PFHxS, further adjusting for recent use of antibiotics did not change effect estimates.

## Discussion

To our knowledge, this is the first study to combine the serum exposome and metabolome using GGMs. The main goal of this study was to explore the links between the chemical exposome and the metabolome and generate hypotheses about possible health effects of exposures to a complex mixture of environmental chemicals. We computed GGMs of non-targeted LC-HRMS data to map direct associations between small molecules. After controlling for multiple testing (FDR < 0.1), we observed many direct associations, including metabolite-metabolite, chemical-metabolite and chemical-chemical associations. We found that most of the significant associations between metabolites corresponded to known biochemical reactions, supporting previous studies on the ability of GGMs to reconstruct metabolic pathways from MS-based metabolomics data^13–15^.

Some metabolite-metabolite associations seem to also reflect molecules originating from the same source of exposures. For example, tryptophan was significantly correlated with arachidonic acid and myristic acid. Although these metabolites are involved in different metabolic pathways, they possibly originated from dietary sources since they can co-occur within the same food component, such as eggs or meat^19^.

When stratifying by occupation, most of metabolite-metabolite associations remained unchanged, while several chemical exposure-metabolite associations were only significant in FF. For example, the association between PFHxS and sulfolithocholylglycine was significant in FF (PCC = 0.12, *p* = 2.6×10^-5^), but not observed in the whole cohort (PCC = 0.05, *p* = 0.01) or in OW (PC C= −0.03, *p* = 0.02). The instability of certain associations is possibly due to inter-occupation variability in the levels of chemical exposures as well as the small sample size. LC-MS/MS analysis of PFAS revealed that serum levels of PFHxS were higher in FF [median = 4.1 ng/mL and interquartile range (IQR) = 5.7 ng/mL] compared to OW (median = 2.5 ng/mL and IQR = 3.8 ng/mL)^16^.

GGMs revealed that certain environmental chemicals were linked to endogenous metabolites involved in cholesterol homeostasis, energy metabolism, inflammation and steroid hormones biosynthesis. We found direct associations between 4-hydroxyactophenone, PFHxS and bile acids, including sulfolithocholylglycine, lithocholic acid glycine and chenodeoxycholic acid (CDCA) in women FF. PFHxS is a perfluoroalkyl substance (PFAS) used as an additive in a wide range of consumer products and food packaging due to surfactant and stain resistant properties^20^. PFAS are also components of firefighting foam, a possible source of exposure for women FF. A previous study of firefighters found positive associations between serum concentrations of PFOS and PFHxS and the number of years using firefighting foam^21^. As previously mentioned, we also observed higher serum PFHxS in women FF, compared to the control group of OW^16^.

Parabens are widely used as antimicrobial preservatives in personal care products, pharmaceuticals, and food and beverages^22,23^. Bile acids (BAs) are cholesterol-derived signaling molecules with hormonal actions that regulate lipid, glucose and energy metabolism by activating several nuclear receptors, including the constitutive androstane receptor (CAR), pregnane X receptor (PXR), farnesoid X receptor (FXR) and vitamin D receptor (VDR), as well as G-protein coupled receptors^18,24,25^. For example, FXR activation has been found to induce expression of peroxisome proliferator activated receptor (PPAR)α and promote oxidation of lipids^26,27^. Previous research suggests that modulation of BAs levels may be an effective strategy for the treatment of components of MetS^28^.

Based on the correlations between PFHxS and BAs in FF, we hypothesized that exposure to PFHxS is associated with MetS in women. When assessing components of MetS in NHANES, we found higher levels of PFHxS to be associated with decreased odds of hyperlipidemia (i.e. HDL cholesterol < 50 mg/dL) in adult women 20-79 years of age enrolled in NHANES. However, PFHxS had neutral effects on the prevalence of the MetS in adult women. A previous study showed that increased levels of serum PFHxS were associated with a lower prevalence of central obesity among male and female adults participants of NHANES 1999-2000 and 2003-2004^29^, but we did not see that association in our NHANES analysis of female adults. Although the study by Lin et al.^29^ found that increased serum PFHxS were correlated with decreased odds of hyperlipidemia, the association did not reach statistical significance. Another study found that early-life exposure to PFHxS was inversely associated with serum concentrations of leptin – a marker of adiposity and metabolic dysfunction – among children^30^. Similar to our results, these studies observed no evidence for an adverse effect of PFHxS on the prevalence of MetS.

BAs also mediate inflammatory and immunomodulatory actions^31,32^. BAs have detergent properties, in that they dissolve fats, and they can cause membrane perturbations, resulting in production of pro-inflammatory molecules and reactive oxygen and nitrogen species which damage DNA, contributing to mutation and apoptosis^33,34^ In addition to finding PFHxS correlated with BAs in our cohort, we also observed significant associations between PFOS, ethyl paraben and butyl paraben, and endogenous molecules involved in inflammatory reactions. Consistent with our GGM findings for PFOS, we found that among adult women in NHANES, serum PFOS concentrations were positively associated with inflammatory and immune reactions (positive associations with CRP and lymphocyte count), while PFHxS was negatively associated with the absolute neutrophil count. Also consistent with our GGM findings, ethyl paraben and butyl paraben were negatively associated with inflammation markers in NHANES. Butyl paraben was also associated with a decrease in white blood cell count, while ethyl paraben did not affect markers of immunity. In previous cross-sectional studies, no significant associations were found between serum PFOS and CRP, and conflicting results were observed for other immune-related health conditions^35^. However, a recent prospective study has observed that elevated PFHxS concentrations at age 7 decreased anti-diphtheria and anti-tetanus antibody concentrations at age 13^36^. Regarding parabens, a previous study among pregnant women found that urinary concentrations of propyl paraben were associated with lower CRP concentrations^37^. Propyl paraben and butyl paraben have also been reported to increase the prevalence of aeroallergen sensitization in children^38,39^.

The underlying mechanisms that mediate the effects of PFAS on cholesterol and glucose homeostasis, inflammation and immune response are thought to be related to PPAR-α,γ activation and fatty oxidation pathways^40-42^. The sub-networks of direct associations found in the present study seem to reinforce these relationships, which may possibly be mediated by PPAR-induced disruption of BAs metabolism and transport. Studies on the association between PFAS and BAs are limited. A previous animal study has demonstrated that mice treated with PFDA resulted in increased serum BAs, probably due to PPAR-α–driven down-regulation of BA transporters involved in the enterohepatic circulation, including the Na+-taurocholate cotransporting polypeptide (NTCP)^43^. Recent studies have shown that PFHxS is a substrate for NTCP in both human and rat hepatocytes^44,45^. A case study involving eight subjects has found that oral treatment with BAs sequestrant increased fecal elimination of PFHxS^46^. Although no human study supporting the paraben-BAs association was found, *in vitro* studies have observed that paraben acts as a strong obesogenic compound through PPAR-γ activation at high concentrations (high μM range) in murine 3T3-L1 cells^47,48^.

Since the BAs associated with PFHxS are produced by intestinal bacteria^34^, we hypothesized that the gut microflora composition may confound associations between these chemicals and effects on metabolic function and inflammation. But adjusting for recent use of antibiotics – as a proxy for loss of bacterial diversity – did not modify the associations. Further studies employing direct measurements of gut bacteria diversity (e.g. targeted measurements of microbial-derived metabolites or 16S rRNA sequencing to characterize microbial communities) are needed to decipher the possible role of the microbiome on the association between chemical exposures and health outcomes.

Many exposure-metabolite associations were not further tested because chemical exposures and/or health outcomes hypothesized were not measured in NHANES. This included direct associations of phthalates with steroid hormones, and metabolites involved in oxidative stress and inflammatory response (Fig. 3A and Table 1). The associations we observed are discussed below in the context of relevant studies from the literature. Phthalates are primarily used as plasticizers in automotive, building materials and consumers products^49^ and known endocrine disrupting compounds. In our study, mono(hydroxyisononyl) phthalate (MHINP) – a major oxidative metabolite of diisononyl phthalate (DINP) – was negatively associated with one steroid hormone metabolite (11β-hydroxyprogesterone). Epidemiological studies have reported that phthalate exposure is associated with decreased levels of steroid hormones in males and females^50-53^. Meeker and Ferguson observed a suggestive negative association between DINP exposure and serum testosterone in women 40-60 years of age enrolled in NHANES^52^. Several phthalates affected steroid hormone synthesis pathways in human adrenocortical carcinoma cell lines (H295R assay), and different phthalates affected different parts of the pathway^54^. Phthalate exposure has also been linked with inflammation and oxidative stress^55-59^. Among phthalate metabolites associated with lipid peroxidation in our study, urinary levels of mono-(2-ethyl)-hexyl phthalate (MEHP) [metabolite of di(2-ethylhexyl) phthalate (DEHP)] have been positively associated with increased serum levels of gamma glutamyltransferase (GGT) – a sensitive biomarker of oxidative stress – while mono-isobutyl phthalate (MiBP) [metabolite of dibutyl phthalate (DBP)] was linked to serum levels of CRP in participants enrolled in NHANES^55^.

This study has several limitations. First, results are correlative in nature and from a small sample size. Second, due to the cross-sectional study design, we are unable to determine whether higher exposures to these chemicals precede changes in levels of endogenous metabolites or are a consequence of metabolic perturbations due to changes in health conditions, diet or exposures to other environmental factors. Third, because no health outcomes were measured in the occupational cohort, exposure-metabolite-outcome associations were tested in another cohort (i.e. NHANES). These two populations probably exhibit significant differences in terms of chemical exposure levels. For example, median serum concentrations of PFHxS in both FF (4.05 ng/mL) and OW (2.55 ng/mL) were twice as high as those measured in NHANES (1.6 ng/mL for cycles 2003-2014)^16^. As such, our NHANES analysis may underestimate or overestimate the associations between chemical exposures and hypothesized health outcomes (e.g. MetS or inflammation) in the occupational cohort. Fourth, due to the LC-HRMS analysis in negative ionization mode, most of endogenous molecules tentatively identified are lipids or hormones. Also, we only identified potential chemicals in the cohort by using an in-house database of 700 environmental chemicals^12^, and so we may have missed other chemical-metabolite interactions that are present in the cohort. We may have missed other important molecules that are more easily detected in positive ionization mode, such as amino acids or vitamins, and involved in other metabolic pathways that would have generated additional exposure-outcome hypotheses.

Despite these limitations, the present study has several strengths. First, this study employed a data-driven approach that combines simultaneous measurements of a wide spectrum of environmental chemicals and endogenous metabolites detected in serum to generate hypotheses related to possible health effects of chemical exposures. This approach provides a comprehensive exploration of the impact of chemicals, individually or in combination, on critical metabolic processes. Second, exposomics and metabolomics data were combined using GGMs that provide identification of direct associations between chemicals and metabolic pathways. Pearson correlation coefficients are generally high in omics data and may lead to identification of many indirect associations that are not biologically relevant. GGMs that are based on partial correlation coefficients provide an estimate of the conditional dependencies between variables and limit the selection of indirect associations. Third, some hypotheses generated from the non-targeted GGM analyses were tested using data from a representative sample of the U.S. population (i.e. NHANES) comprising adult women from diverse racial/ethnic background and socio-economic status, and some of these findings were consistent. For example, analysis of NHANES data confirmed associations found in GGMs between exposures to PFHxS and PFOS and cholesterol metabolism and inflammation. Fourth, GGMs revealed that exposures to previously unstudied chemicals (e.g. 4-hexyloxyphenol or certain phthalates metabolites) may potentially have adverse effects on critical metabolic pathways such as metabolism of arachidonic acid and linoleic acid involved in lipid peroxidation and inflammatory and immune responses as well as steroid hormone biosynthesis. Taken together, our findings demonstrate a novel approach to discovering associations between chemical exposures and upstream biological processes potentially involved in disease. We encourage further studies to apply these techniques in other cohorts and to further evaluate associations between chemical exposures and health outcomes that we have reported here.

## Data Availability

The data that support the findings of this study are available from the corresponding authors on request

## METHODS

### Study population

This study was conducted using LC-QTOF/MS data available from an established cohort of California women workers known as the Women Firefighter Biomonitoring Collaborative (WFBC). The WFBC is a cross-sectional study designed to measure and compare exposures to potential breast carcinogens and other endocrine disrupting compounds (EDCs) in 69 San Francisco (SF) women firefighters (FF) and 74 female controls among office workers (OW) from the City of San Francisco. Detailed description of this cohort has been published elsewhere^12,16^. Demographic characteristics and breast cancer risk factor information, including menopausal status, history of hormone replacement therapy, and reproductive history were collected using questionnaires during in-person interviews. Body Mass Index (BMI; kg. m^-2^) was calculated from participants’ height and weight measured at the time of the in-person interview. A 50 mL blood sample was collected by a certified phlebotomist. All participants were consented into the study following protocols approved by the Institutional Review Board of the University of California, Berkeley (# 2013-07-5512).

Overall, the FF and OW cohorts were similar in terms of age, race/ethnicity, BMI, parity, and hormone use (Table 1). However, the household income for women FF was significantly higher compared to OW. There were significantly more pre-menopausal women in the FF group and women FF had a higher proportion of college graduates but fewer graduate degrees than the OW.

### Non-targeted LC-QTOF/MS metabolomics analysis

Non-targeted analysis of serum samples was performed as previously described^60^. Briefly, 250 μL of serum sample was spiked with 2.5 μL of 1 μg/mL of internal standard (2.5 ng BPA-d16) and centrifuged at 3,000 rpm for 10 min. Analytes were extracted using solid-phase extraction (SPE; Waters Oasis HLB 10 mg, 1cc). Extracts were dried under a stream of nitrogen gas and reconstituted in 250 μL of 10% methanol.

Extracts were analyzed on a LC-QTOF/MS system consisting of an LC 1260 autosampler, pumps and a QTOF/MS 6550 (Agilent, Santa Cruz, CA, USA). Analytes were separated by a reversed-phase method using a C18 column (Agilent Poroshell 120, 2.1 mm × 100 mm, 2.7 μm particle size) maintained at 55°C. Mobile phase A consisted of water with 0.05% ammonium acetate (pH=7.8) and mobile phase B consisted of methanol with 0.05% ammonium acetate (pH=7.8). The elution gradient employed was: 0-0.5 min, 5% B; 1.5 min, 30% B; 4.5 min, 70% B; 7.5-10 min, 100% B; 10.01-14 min, 5% B. The injection volume was 50 μL. Analyses were performed with a QTOF/MS operating in negative electrospray ionization mode (ESI-). Ions were collected in the m/z 80–600 range at high resolution for eluates coming out of the LC from 1-12 min. Using the Auto MS/MS mode (information-dependent acquisition), a product ion scan (MS/MS) of the three most abundant peaks at high resolution was triggered each time a precursor ion with an intensity of ≥500 counts/second was generated in the TOF-MS scan using a collision voltage ranging from 0 to 40 V depending of ions m/z. The LC-QTOF/MS analysis produces a total ion chromatogram for each sample, which includes the following: the accurate mass of each unique compound (expressed as *m/z* of their respective anion), peak area, retention time and spectral data on the parent and fragment ions, including isotopic pattern.

### Data processing

#### Exposome annotation

Chemical exposures were identified from non-targeted LC-QTOF/MS data as previously described^12,60^. Briefly, all detected *m/z* were matched with those from an in-house MS database of environmental chemicals with a mass tolerance value of 10 ppm. The in-house database consists of more 700 chemicals: 1) environmental organic acids including parabens and paraben metabolites, phthalates and phthalate metabolites and pesticides and pesticide metabolites; 2) chemicals that increase breast cancer risk including mammary carcinogens and mammary gland developmental disruptors^61,62^; 3) known firefighting-related occupational exposures including perfluorinated compounds found in firefighting foams, polychlorinated and polybrominated dioxins and furans and other flame retardants. A list of tentatively annotated chemicals was generated for all samples, with corresponding exact mass, retention time, mass error, peak area, chemical formula, and match scores. Then, we performed retention time correction using an inhouse R script in order to align LC-QTOF/MS data. We assigned the chemicals with the same formula to be two different entities if the difference in retention time of two adjacent chemicals was greater than 0.16 min. The exposome data matrix consisted of peak area and *m/z* of 620 unique chemicals which matched to 300 chemical formulas^12^.

#### Metabolome annotation

LC-QTOF/MS metabolomics data were pre-processed using the R package “XCMS”^63^ for peak detection, retention time correction and peak alignment (the R script used for data pre-processing can be found in Supplementary Information). After pre-processing, a data matrix containing retention time, mass-to-charge ratio *(m/z)* and intensity of features was generated. Metabolomics features were annotated using the R package “xMSannotator”^64^. We used the Human Metabolome Database 3.6 (HMDB) that contains detailed molecular information about 42,632 small molecules^65^ as the reference library for annotation of metabolomics features. We retained only annotated molecules with a confidence score equal to 3 (the highest score of confidence) and classified by HMDB as previously detected and quantified in biological matrices (the R script used for annotation of metabolomics features can be found in Supplementary Information). After this process, the metabolome data matrix contained *m/z* and intensity of 90 annotated molecules (Supplementary Table S5).

The identification level of environmental chemicals and endogenous metabolites was reported as proposed by the Metabolic Standards Initiative^17^: level 1: match based on accurate mass (± 10 ppm), fragmentation pattern and relative retention time with authentic standards; level 2: match based on accurate mass (± 10 ppm) and fragmentation pattern using mass spectra from public metabolomics libraries or in-silico fragmentation software^66^.; level 3: match based on accurate mass (± 10 ppm) using public metabolomics libraries. Most small molecules tentatively identified in this study corresponded to level 2 or level 3 annotation. Eight environmental chemicals were validated as level 1 annotation (Supporting Information Table S1).

### Statistical analysis

#### Gaussian graphical modeling (GGM)

GGMs were built by combining the exposome data matrix with the metabolome data matrix. We first excluded molecules with more than 50% missing values. The filtered data matrix contained 69 samples × 145 molecules (55 environmental chemicals and 90 endogenous metabolites), 74 samples × 139 molecules (49 environmental chemicals and 90 endogenous metabolites), and 143 samples × 142 molecules (52 environmental chemicals and 90 endogenous metabolites) for women FF, OW and the whole cohort, respectively. Missing values imputation, data normalization and transformation were conducted using MetaboAnalyst 3.0^67^. Remaining missing values were imputed using the k-nearest neighbor (KNN) method^68^. Exposome data (i.e. peak area) and metabolome data (i.e. metabolite intensity) were normalized, separately, by sum and generalized log-transformed to reduce analytical variations. We computed GGMs using the ggm.estimator.pcor function from the R package “GeneNet”^69^. We considered partial correlations between two molecules to be significant if the resulting *p* value was below the False Discovery Rate (FDR) threshold of 0.1 (for FF *p* < 1.79×10^-4^, for OW *p* < 1.21×10^-4^ and for the whole cohort *p* < 1.71×10^-4^). The GGM networks were constructed and visualized with Cytoscape 3.8.0^70^ using “organic” as layout. Edges connecting nodes represent significant partial correlations. Partial correlation coefficients were used as the edge attributes.

#### Validation of exposome-metabolome associations

For each significant exposure-metabolite partial correlation, we used several approaches to attempt to validate the associations by checking whether a similar type of chemical-effect relationship has been reported elsewhere, for example in National Health and Nutrition Examination Survey (NHANES) or in PubMed.

We used NHANES to confirm or refute the association found in our occupational cohort if 1) the chemical of interest and 2) one marker of the biological pathway perturbed were measured in at least one NHANES cycle. NHANES is an ongoing cross-sectional study of the civilian noninstitutionalized U.S. population designed to collect data on dietary and health factors (ref, see Supplementary Information for more details). NHANES study protocols were approved by the National Center for Health Statistics’ Research Ethics Review Board. Since our study population consists of women workers, we selected only non-pregnant women adults (20-79 years of age) enrolled in NHANES. All NHANES data (demographics, examination data, laboratory data, and questionnaire data) were downloaded using the R package “RNHANES” ^71^. Full description of exposure measurements, outcomes and covariates in NHANES can be found in Supplementary Information. To test our hypothesis that certain environmental chemicals measured in women’s serum affect the level of inflammatory signaling molecules, we explored the relationships between serum or urinary concentrations of each chemical and serum markers of inflammation [C-reactive protein (CRP), and the complete blood count which measured the number of white blood cells, lymphocytes, neutrophils, monocytes, eosinophils, basophils and platelets] in adult women 20-79 years of age. Details about measurements of environmental chemicals and markers of inflammation in NHANES can be found in Supplementary Information. Since we were interested only in the effects of chemicals on chronic inflammation, we excluded participants who reported poor health status, or acute infection at the time of examination, including head or chests colds, stomach or intestinal illness, or flu, pneumonia, or ear infection. We further excluded individuals with CRP concentrations > 10 mg/L because these extreme values likely reflect acute inflammation ^72,73^. We constructed multivariable linear regression models with natural log-transformed CRP or absolute blood cells count as the dependent variable and one natural log-transformed chemical divided by 100 as a predictor (e.g. [ln(PFOS)]/100).

To test the hypothesis that exposures to PFHxS are associated with metabolic syndrome (MetS) through alteration of bile acid metabolism, we evaluated the relationships between serum PFHxS and MetS in women adults 20-79 years of age. Data were pooled from NHANES 2003-2014 for PFHxS. We excluded participants with fasting time <8 hours. We used the National Cholesterol Education Program’s Adult Treatment Panel III report (NCEP/ATPIII) to define MetS ^74^. The NCEP/ATPIII classifies women as having MetS, if at least 3 of the following 5 criteria are met: 1) waist circumference ≥ 88 cm; 2) triglyceride ≥ 150 mg/dL; 3) high density lipid (HDL) cholesterol < 50 mg/dL; 4) blood pressure ≥ 130/85 mmHg or treatment for hypertension; 5) fasting blood glucose ≥ 100 mg/dL or treatment for diabetes (further details can be found in Supplementary Information). To assess associations between chemical measurements and MetS, we used logistic regression models to estimate adjusted odds ratio (ORs) and corresponding 95% CIs. Serum concentrations of PFHxS were natural log-transformed to address skewness.

All models were adjusted for likely sources of confounding, including age (years, continuous), race and ethnicity (non-Hispanic white, non-Hispanic black, Mexican American, multiracial or other), and poverty/income ratio [PIR (the ratio of self-reported family income to the family’s appropriate threshold value), divided into tertiles]. When urinary concentrations of chemicals were used as predictors, we further adjusted for urinary creatinine (continuous, natural log-transformed) to adjust for urine dilution. For models with inflammatory markers as dependent variables, we further adjusted for serum cotinine (natural log-transformed, continuous exposure) as a measure of exposure to tobacco smoke. For models with MetS and individual components of MetS as dependent variables, we further adjusted for self-reported physical activity (none, moderate or vigorous), smoking status (never, past or current), and total caloric intake derived from the average of two-day 24 recalls and divided into quartiles. Association estimates for models with non-persistent chemicals (i.e. ethyl paraben or propyl paraben) as predictors were reported before and after adjustment for body mass index (BMI) since it can be argued that BMI can act as a confounder or a mediator. For chemicals associated with bile acids in GGM networks (i.e. PFHxS), ORs and association estimates were also reported before and after further adjustment for recent use of antibiotics (in the past 30 days, dichotomous) since the gut microbiota is a possible confounder of associations between exposures to these chemicals and outcome variables related to MetS and inflammation. Data were analyzed using the R package “survey” to obtain estimates of association or ORs and 95% CIs accounting for the complex NHANES sampling design. We also used the weights to adjust for the oversampling of certain population subgroups and to account for non-response and non-coverage in NHANES. When multiple NHANES cycles were combined, we recalculated new sample weights for each participant by dividing the 2-year sample weights provided by the number of cycles combined. All tests were two sided, and *p* < 0.05 was the level of significance.

## Acknowledgements

The authors thank all of the WFBC study participants for their contribution to the study. This work is supported by the California Breast Cancer Research Program #19BB-2900 (RG, VB, RRG, JT, TL, HB, RAR, RMF), the National Institute of Environmental Health Sciences R01ES027051 (RMF),the San Francisco Firefighter Cancer Prevention Foundation (HB), and the Silent Spring Institute Innovation Fund (RAR and VB). We thank Anthony Stefani, Emily O’Rourke, Nancy Carmona, Karen Kerr, Julie Mau, Natasha Parks, Lisa Holdcroft, former SF Fire Chief Joanne Hayes-White, SF Fire Chief Jeanine Nicholson, Sharyle Patton, Connie Engel and Nancy Buermeyer for their contributions to the study.

## Author contributions

RAR, RMF, RRG and HB designed the WFBC study. RRG and TL performed non-targeted analysis of serum samples. RRG, RAR and RMF developed the in-house MS database of environmental chemicals used to annotate the serum exposome. RRG, TL, RG and VB generated the serum exposome data. VB generated the serum metabolome data, designed and led the data analysis to combine the exposome and metabolome data, and wrote the initial draft of the manuscript. JT, RMF, RAR and RRG edited the submitted version. All authors critically revised the draft and approved the final manuscript.

## References

1. Rappaport, S. M. Genetic Factors Are Not the Major Causes of Chronic Diseases. PloS One 11, e0154387 (2016).

2. Wild, C. P. Complementing the genome with an ‘exposome’: the outstanding challenge of environmental exposure measurement in molecular epidemiology. Cancer Epidemiol. Biomark. Prev. Publ. Am. Assoc. Cancer Res. Cosponsored Am. Soc. Prev. Oncol. 14, 1847–1850 (2005).

3. Bessonneau, V., Pawliszyn, J. & Rappaport, S. M. The Saliva Exposome for Monitoring of Individuals’ Health Trajectories. Environ. Health Perspect. 125, 077014 (2017).

4. German, J. B., Hammock, B. D. & Watkins, S. M. Metabolomics: building on a century of biochemistry to guide human health. Metabolomics Off J. Metabolomic Soc. 1, 3–9 (2005).

5. Nicholson, J. K. & Wilson, I. D. Opinion: understanding ‘global’ systems biology: metabonomics and the continuum of metabolism. Nat. Rev. Drug Discov. 2, 668–676 (2003).

6. Koeth, R. A. et al. Intestinal microbiota metabolism of L-carnitine, a nutrient in red meat, promotes atherosclerosis. Nat. Med. 19, 576–585 (2013).

7. Tang, W. H. W. et al. Intestinal Microbial Metabolism of Phosphatidylcholine and Cardiovascular Risk. http://dx.doi.org/10.1056/NEJMoa1109400 http://www.nejm.org/doi/10.1056/NEJMoa1109400 (2013) doi:10.1056/NEJMoa1109400.

8. Wang, Z. et al. Gut flora metabolism of phosphatidylcholine promotes cardiovascular disease. Nature 472, 57–63 (2011).

9. Patel, C. J., Bhattacharya, J. & Butte, A. J. An Environment-Wide Association Study (EWAS) on Type 2 Diabetes Mellitus. PLOS ONE 5, e10746 (2010).

10. Tzoulaki, I. et al. A nutrient-wide association study on blood pressure. Circulation 126, 2456–2464 (2012).

11. Patel, C. J. et al. Systematic evaluation of environmental and behavioural factors associated with all-cause mortality in the United States national health and nutrition examination survey. Int. J. Epidemiol. 42, 1795–1810 (2013).

12. Grashow, R. et al. Integrating exposure knowledge and serum suspect screening as a new approach to biomonitoring: An application in firefighters and office workers. Environ. Sci. Technol. (2020) doi:10.1021/acs.est.9b04579.

13. Krumsiek, J., Suhre, K., Illig, T., Adamski, J. & Theis, F. J. Gaussian graphical modeling reconstructs pathway reactions from high-throughput metabolomics data. BMC Syst. Biol. 5, 21 (2011).

14. Jourdan, C. et al. Body fat free mass is associated with the serum metabolite profile in a population-based study. PloS One 7, e40009 (2012).

15. Mittelstrass, K. et al. Discovery of sexual dimorphisms in metabolic and genetic biomarkers. PLoS Genet. 7, e1002215 (2011).

16. Trowbridge, J. et al. Exposure to Perfluoroalkyl Substances in a Cohort of Women Firefighters and Office Workers in San Francisco. Environ. Sci. Technol. (2020) doi:10.1021/acs.est.9b05490.

17. Sumner, L. W. et al. Proposed minimum reporting standards for chemical analysis Chemical Analysis Working Group (CAWG) Metabolomics Standards Initiative (MSI). Metabolomics Off. J. Metabolomic Soc. 3, 211–221 (2007).

18. Chiang, J. Y. L. Bile acids: regulation of synthesis. J. Lipid Res. 50, 1955–1966 (2009).

19. USDA. USDA Food Composition Databases. (2018).

20. Björklund, J. A., Thuresson, K. & De Wit, C. A. Perfluoroalkyl compounds (PFCs) in indoor dust: concentrations, human exposure estimates, and sources. Environ. Sci. Technol. 43, 2276–2281 (2009).

21. Rotander, A., Toms, L.-M. L., Aylward, L., Kay, M. & Mueller, J. F. Elevated levels of PFOS and PFHxS in firefighters exposed to aqueous film forming foam (AFFF). Environ. Int. 82, 28–34 (2015).

22. Calafat, A. M., Ye, X., Wong, L.-Y., Bishop, A. M. & Needham, L. L. Urinary Concentrations of Four Parabens in the U.S. Population: NHANES 2005-2006. Environ. Health Perspect. 118, 679–685 (2010).

23. Dodson, R. E. et al. Endocrine Disruptors and Asthma-Associated Chemicals in Consumer Products. Environ. Health Perspect. 120, 935–943 (2012).

24. Chiang, J. Y. L. Bile acid metabolism and signaling. Compr. Physiol. 3, 1191–1212 (2013).

25. Kliewer, S. A. & Mangelsdorf, D. J. Bile Acids as Hormones: The FXR-FGF15/19 Pathway. Dig. Dis. Basel Switz. 33, 327–331 (2015).

26. Pineda Torra, I. et al. Bile acids induce the expression of the human peroxisome proliferator-activated receptor alpha gene via activation of the farnesoid X receptor. Mol. Endocrinol. Baltim. Md 17, 259–272 (2003).

27. Zhang, Y., Castellani, L. W., Sinal, C. J., Gonzalez, F. J. & Edwards, P. A. Peroxisome proliferator-activated receptor-γ coactivator 1α (PGC-1α) regulates triglyceride metabolism by activation of the nuclear receptor FXR. Genes Dev. 18, 157–169 (2004).

28. Ma, H. & Patti, M. E. Bile acids, obesity, and the metabolic syndrome. Best Pract. Res. Clin. Gastroenterol. 28, 573–583 (2014).

29. Lin, C.-Y., Chen, P.-C., Lin, Y.-C. & Lin, L.-Y. Association Among Serum Perfluoroalkyl Chemicals, Glucose Homeostasis, and Metabolic Syndrome in Adolescents and Adults. Diabetes Care 32, 702–707 (2009).

30. Fleisch, A. F. et al. Early-Life Exposure to Perfluoroalkyl Substances and Childhood Metabolic Function. Environ. Health Perspect. 125, 481–487 (2017).

31. Mencarelli, A. et al. The bile acid sensor farnesoid X receptor is a modulator of liver immunity in a rodent model of acute hepatitis. J. Immunol. Baltim. Md 1950 183, 6657–6666 (2009).

32. Vavassori, P., Mencarelli, A., Renga, B., Distrutti, E. & Fiorucci, S. The bile acid receptor FXR is a modulator of intestinal innate immunity. J. Immunol. Baltim. Md 1950 183, 6251–6261 (2009).

33. Ding, L., Yang, L., Wang, Z. & Huang, W. Bile acid nuclear receptor FXR and digestive system diseases. Acta Pharm. Sin. B 5, 135–144 (2015).

34. Ridlon, J. M. & Bajaj, J. S. The human gut sterolbiome: bile acid-microbiome endocrine aspects and therapeutics. Acta Pharm. Sin. B 5, 99–105 (2015).

35. Chang, E. T., Adami, H.-O., Boffetta, P., Wedner, H. J. & Mandel, J. S. A critical review of perfluorooctanoate and perfluorooctanesulfonate exposure and immunological health conditions in humans. Crit. Rev. Toxicol. 46, 279–331 (2016).

36. Grandjean, P. et al. Serum Vaccine Antibody Concentrations in Adolescents Exposed to Perfluorinated Compounds. Environ. Health Perspect. 125, 077018 (2017).

37. Watkins, D. J. et al. Associations between urinary phenol and paraben concentrations and markers of oxidative stress and inflammation among pregnant women in Puerto Rico. Int. J. Hyg. Environ. Health 218, 212–219 (2015).

38. Savage, J. H., Matsui, E. C., Wood, R. A. & Keet, C. A. Urinary levels of triclosan and parabens are associated with aeroallergen and food sensitization. J. Allergy Clin. Immunol. 130, 453-60.e7 (2012).

39. Spanier, A. J., Fausnight, T., Camacho, T. F. & Braun, J. M. The associations of triclosan and paraben exposure with allergen sensitization and wheeze in children. Allergy Asthma Proc. 35, 475–481 (2014).

40. Buhrke, T., Kibellus, A. & Lampen, A. In vitro toxicological characterization of perfluorinated carboxylic acids with different carbon chain lengths. Toxicol. Lett. 218, 97–104 (2013).

41. Wolf, C. J., Schmid, J. E., Lau, C. & Abbott, B. D. Activation of mouse and human peroxisome proliferator-activated receptor-alpha (PPARα) by perfluoroalkyl acids (PFAAs): further investigation of C4-C12 compounds. Reprod. Toxicol. Elmsford N 33, 546–551 (2012).

42. Zhang, L., Ren, X.-M., Wan, B. & Guo, L.-H. Structure-dependent binding and activation of perfluorinated compounds on human peroxisome proliferator-activated receptor γ. Toxicol. Appl. Pharmacol. 279, 275–283 (2014).

43. Cd, C. X. and K. Critical role of PPAR-alpha in perfluorooctanoic acid- and perfluorodecanoic acid-induced downregulation of Oatp uptake transporters in mouse livers. – PubMed – NCBI. https://www.ncbi.nlm.nih.gov/pubmed/18703564.

44. Han, X., Nabb, D. L., Russell, M. H., Kennedy, G. L. & Rickard, R. W. Renal elimination of perfluorocarboxylates (PFCAs). Chem. Res. Toxicol. 25, 35–46 (2012).

45. Zhao, W. et al. Na+/Taurocholate Cotransporting Polypeptide and Apical Sodium-Dependent Bile Acid Transporter Are Involved in the Disposition of Perfluoroalkyl Sulfonates in Humans and Rats. Toxicol. Sci. Off. J. Soc. Toxicol. 146, 363–373 (2015).

46. Genuis, S. J., Curtis, L. & Birkholz, D. Gastrointestinal Elimination of Perfluorinated Compounds Using Cholestyramine and Chlorella pyrenoidosa. International Scholarly Research Notices https://www.hindawi.com/journals/isrn/2013/657849/ (2013) doi:10.1155/2013/657849.

47. Hu, P. et al. Effects of Parabens on Adipocyte Differentiation. Toxicol. Sci. 131, 56–70 (2013).

48. Pereira-Fernandes, A. et al. Evaluation of a Screening System for Obesogenic Compounds: Screening of Endocrine Disrupting Compounds and Evaluation of the PPAR Dependency of the Effect. PLoS ONE 8, (2013).

49. Giovanoulis, G. et al. Multi-pathway human exposure assessment of phthalate esters and DINCH. Environ. Int. 112, 115–126 (2018).

50. Hauser, R. et al. Male reproductive disorders, diseases, and costs of exposure to endocrine-disrupting chemicals in the European Union. J. Clin. Endocrinol. Metab. 100, 1267–1277 (2015).

51. Marsee, K., Woodruff, T. J., Axelrad, D. A., Calafat, A. M. & Swan, S. H. Estimated daily phthalate exposures in a population of mothers of male infants exhibiting reduced anogenital distance. Environ. Health Perspect. 114, 805–809 (2006).

52. Meeker, J. D. & Ferguson, K. K. Urinary Phthalate Metabolites Are Associated With Decreased Serum Testosterone in Men, Women, and Children From NHANES 2011–2012. J. Clin. Endocrinol. Metab. 99, 4346–4352 (2014).

53. Mendiola, J. et al. Urinary concentrations of di(2-ethylhexyl) phthalate metabolites and serum reproductive hormones: pooled analysis of fertile and infertile men. J. Androl. 33, 488–498 (2012).

54. Haggard, D. E. et al. High-Throughput H295R Steroidogenesis Assay: Utility as an Alternative and a Statistical Approach to Characterize Effects on Steroidogenesis. Toxicol. Sci. Off. J. Soc. Toxicol. 162, 509–534 (2018).

55. Ferguson, K. K., Loch-Caruso, R. & Meeker, J. D. Urinary Phthalate Metabolites in Relation to Biomarkers of Inflammation and Oxidative Stress: NHANES 1999-2006. Environ. Res. 111, 718–726 (2011).

56. Ferguson, K. K., Loch-Caruso, R. & Meeker, J. D. Exploration of oxidative stress and inflammatory markers in relation to urinary phthalate metabolites: NHANES 1999–2006. Environ. Sci. Technol. 46, 477–485 (2012).

57. Ferguson, K. K., McElrath, T. F., Chen, Y.-H., Mukherjee, B. & Meeker, J. D. Urinary Phthalate Metabolites and Biomarkers of Oxidative Stress in Pregnant Women: A Repeated Measures Analysis. Environ. Health Perspect. 123, 210–216 (2015).

58. Ferguson, K. K. et al. Urinary Phthalate Metabolite Associations with Biomarkers of Inflammation and Oxidative Stress Across Pregnancy in Puerto Rico. Environ. Sci. Technol. 48, 7018–7025 (2014).

59. Guo, Y. et al. Urinary Concentrations of Phthalates in Couples Planning Pregnancy and Its Association with 8-Hydroxy-2′-deoxyguanosine, a Biomarker of Oxidative Stress: Longitudinal Investigation of Fertility and the Environment Study. Environ. Sci. Technol. 48, 9804–9811 (2014).

60. Gerona, R. R. et al. Suspect screening of maternal serum to identify new environmental chemical biomonitoring targets using liquid chromatography-quadrupole time-of-flight mass spectrometry. J. Expo. Sci. Environ. Epidemiol. 28, 101–108 (2018).

61. Rudel, R. A., Attfield, K. R., Schifano, J. N. & Brody, J. G. Chemicals causing mammary gland tumors in animals signal new directions for epidemiology, chemicals testing, and risk assessment for breast cancer prevention. Cancer 109, 2635–2666 (2007).

62. Rudel, R. A., Ackerman, J. M., Attfield, K. R. & Brody, J. G. New exposure biomarkers as tools for breast cancer epidemiology, biomonitoring, and prevention: a systematic approach based on animal evidence. Environ. Health Perspect. 122, 881–895 (2014).

63. Smith, C. A., Want, E. J., O’Maille, G., Abagyan, R. & Siuzdak, G. XCMS: Processing Mass Spectrometry Data for Metabolite Profiling Using Nonlinear Peak Alignment, Matching, and Identification. Anal. Chem. 78, 779–787 (2006).

64. Uppal, K., Walker, D. I. & Jones, D. P. xMSannotator: An R Package for Network-Based Annotation of High-Resolution Metabolomics Data. Anal. Chem. 89, 1063–1067 (2017).

65. Wishart, D. S. et al. HMDB 3.0—The Human Metabolome Database in 2013. Nucleic Acids Res. 41, D801–D807 (2013).

66. Dührkop, K., Shen, H., Meusel, M., Rousu, J. & Böcker, S. Searching molecular structure databases with tandem mass spectra using CSI:FingerID. Proc. Natl. Acad. Sci. U. S. A. 112, 12580–12585 (2015).

67. Xia, J. & Wishart, D. S. Using MetaboAnalyst 3.0 for Comprehensive Metabolomics Data Analysis. in Current Protocols in Bioinformatics (John Wiley & Sons, Inc., 2002). doi:10.1002/cpbi.11.

68. Xia, J. & Wishart, D. S. Web-based inference of biological patterns, functions and pathways from metabolomic data using MetaboAnalyst. Nat. Protoc. 6, 743–760 (2011).

69. Schäfer, J. & Strimmer, K. An empirical Bayes approach to inferring large-scale gene association networks. Bioinformatics 21, 754–764 (2005).

70. Shannon, P. et al. Cytoscape: a software environment for integrated models of biomolecular interaction networks. Genome Res. 13, 2498–2504 (2003).

71. Susmann, H. RNHANES: Facilitates Analysis of CDC NHANES Data. (2016).

72. Shaheen, M. et al. Hepatitis C, metabolic syndrome, and inflammatory markers: results from the Third National Health and Nutrition Examination Survey [NHANES III]. Diabetes Res. Clin. Pract. 75, 320–326 (2007).

73. Yoon, S. S., Dillon, C. F., Carroll, M., Illoh, K. & Ostchega, Y. Effects of statins on serum inflammatory markers: the U.S. National Health and Nutrition Examination Survey 1999-2004. J. Atheroscler. Thromb. 17, 1176–1182 (2010).

74. Grundy, S. M. et al. Definition of metabolic syndrome: report of the National Heart, Lung, and Blood Institute/American Heart Association conference on scientific issues related to definition. Arterioscler. Thromb. Vasc. Biol. 24, e13-18 (2004).

